# Residential Ozone and Risk of Chronic Obstructive Pulmonary Disease in the United States: Demographic Differences in the All of Us Research Program

**DOI:** 10.1101/2025.05.13.25327336

**Authors:** Shelton Lo, Gabriel A. Goodney, Hantao Wang, Jungeun Lim, Susan Valerie Czach, Jared A. Fisher, Joseph J. Shearer, Maryam Hashemian, Véronique L. Roger, Rena R. Jones, Jason Y.Y. Wong

## Abstract

**Background:** The understanding of the complex interactions among ozone, temperature, various atmospheric conditions, and the intricate mixtures that make up the air exposome remains limited. Whether exposure to ozone concentrations consistently below current U.S. regulatory limits (70 ppb, 8-hour average) is associated with newly-diagnosed chronic obstructive pulmonary disease (COPD) is unclear, especially among demographic subgroups. We examined associations between residential ozone and incident COPD in a large prospective cohort study in the U.S., and assessed heterogeneity by demographic subgroups.

**Methods:** The All of Us research program followed >848,000 volunteers enrolled in 2017–2023. Among 596, 926 participants whom consented to the release of EHR data, annual average ozone concentrations from satellite measurements from 2000—2016 were linked to residential location for 376,535 participants. Multivariable Cox regression was used to estimate associations between ozone and incident COPD over the study period, adjusting for co-exposures and potential confounders. We assessed effect modification using cross-product terms and stratified analyses with race, sex, age, income, and smoking status. To account for non-probabilistic sampling, we applied cell weighting based on the distribution of key factors from the 2018—2022 American Community Survey.

**Results:** We identified 7,907 incident COPD cases over an average 4-year follow-up. Residential ozone concentrations (Median: 37.75 ppb, Min-Max: 29.79–51.39) were similar across subgroups. Overall, we observed a positive, non-monotonic relationship between ozone and COPD risk, driven by the highest quartile of exposure (HR_Quartile 4vs1_=1.09, 95% CI: 1.01, 1.18). We found evidence of heterogeneity among different subgroups. Importantly, the exposure-response relationship was monotonic and most apparent in male never-smokers (P-trend_continuous_=6.62×10^-6^).

**Conclusion:** Even consistently below U.S. regulatory limits, the respiratory effects of ozone were apparent independent of smoking. Our findings provide greater precision into which subgroups might be more susceptible.

## Introduction

Chronic obstructive pulmonary disease (COPD) is a progressive condition caused by abnormalities of the airways and/or alveoli that result in persistent airflow obstruction ^1^. COPD is a substantial burden to the people and healthcare systems of the U.S. Although recent reports indicated a slight decline in the incidence rates of COPD among adults aged >50 years, it remains a substantial contributor to morbidity and mortality ^2^ ^3^. In recent years, COPD was the sixth leading cause of death, with over 16 million people diagnosed and 150,000 deaths annually ^2^ ^3^. An aging population contributes to the burden of COPD, largely due to the cumulative effects of long-term environmental exposures and other risk factors over the life course ^4^. While smoking remains the primary risk factor for COPD, environmental exposures, including chemical compounds in the air ^5^, continue to play a significant role in disease risk among both smokers and never-smokers ^6^ ^7^ ^8^.

In the U.S., COPD rates are higher among certain racial and ethnic populations, working-class individuals of lower socioeconomic status (SES), and those living in areas with high environmental burdens ^9^ ^10^ ^11^. Evidence from population-based studies highlight the role of ubiquitous outdoor air pollutants, such as particulate matter 2.5 (PM_2.5_) and nitrogen dioxide (NO_2_), in contributing to COPD risk ^12^. Although studies have reported associations between ozone exposure and COPD ^13^ ^14^, the findings have been inconsistent. There is limited understanding of the complex interrelationships between ozone, temperature, other atmospheric conditions, and complex mixtures of the air exposome. However, it is known that ozone and NO_2_ are closely linked through photochemical reactions, whereas NO_2_ undergoes photodissociation in sunlight to form ozone. Further, whether ozone exposure consistently below the current U.S. National Ambient Air Quality Standard (70 ppb averaged over 8 hours) is associated with increased COPD risk, particularly among demographic subgroups defined by race and ethnicity, sex, age, SES, and smoking remains unclear.

To address these knowledge gaps, we leveraged data from the All of Us (AoU) research program ^15^, a prospective mega cohort ^16^ ^17^ of >848,000 people from across the U.S., to investigate the associations between ozone, other air pollutants, temperature variation, and risk of incident COPD, with attention to characterizing heterogeneity across demographic subgroups. We sought to clarify the complexities surrounding the respiratory effects of ozone in the U.S. population and provide greater precision into which demographic subgroups might be more susceptible, which could contribute to risk stratification analyses and inform targeted population interventions.

## Methods

### Study Population

The AoU Research Program has been previously described in detail ^15^. Briefly, AoU is a prospective cohort study that enrolled over 848,000 volunteer participants from across the U.S. who were ≥18 years of age. Among these participants, 633,547 subjects who met inclusion criteria (≥18 years of age, reside in the United States or its territories, and able to provide informed consent) were prospectively followed between 2017 and 2023 (allofus.nih.gov). Demographic data, including date of birth, sex at birth, race, Hispanic status, education level, annual household income, and smoking status, were obtained from questionnaires. Health and medical information from the >50 affiliated health care provider organizations were obtained from electronic health records (EHRs) standardized under the Observational Medical Outcomes Partnership (OMOP) Common Data Model. Among the participants who met the study’s inclusion criteria, 596,926 participants consented to the release of EHR data. Geolocation data was derived from participant provided addresses or EHR data. Residential history was collected at enrollment through *The Basics* survey questionnaire. Controlled Tier residential geolocation data access is limited to three-digit zip codes to ensure participant privacy and prevent re-identification. Postal regions with <20,000 participants are aggregated into a nearby three-digit zip code. After applying exclusion criteria—including 181,152 participants without EHR data, 22,178 participants with prevalent COPD, and 17,061 participants without valid residential address history—the final analytic sample comprised 376,535 individuals. Annual average ozone concentrations derived from satellite-based measurements (2000–2016) were linked to residential addresses for these 376,535 individuals. Figure 1 details how the analytical sample was obtained. Written informed consent was obtained from all participants. All data were fully anonymized before researcher access and data use. All data analysis was conducted using the latest AoU Controlled Tier Dataset v8 (C2024Q3R4) released in February 2025.

**Figure 1:**
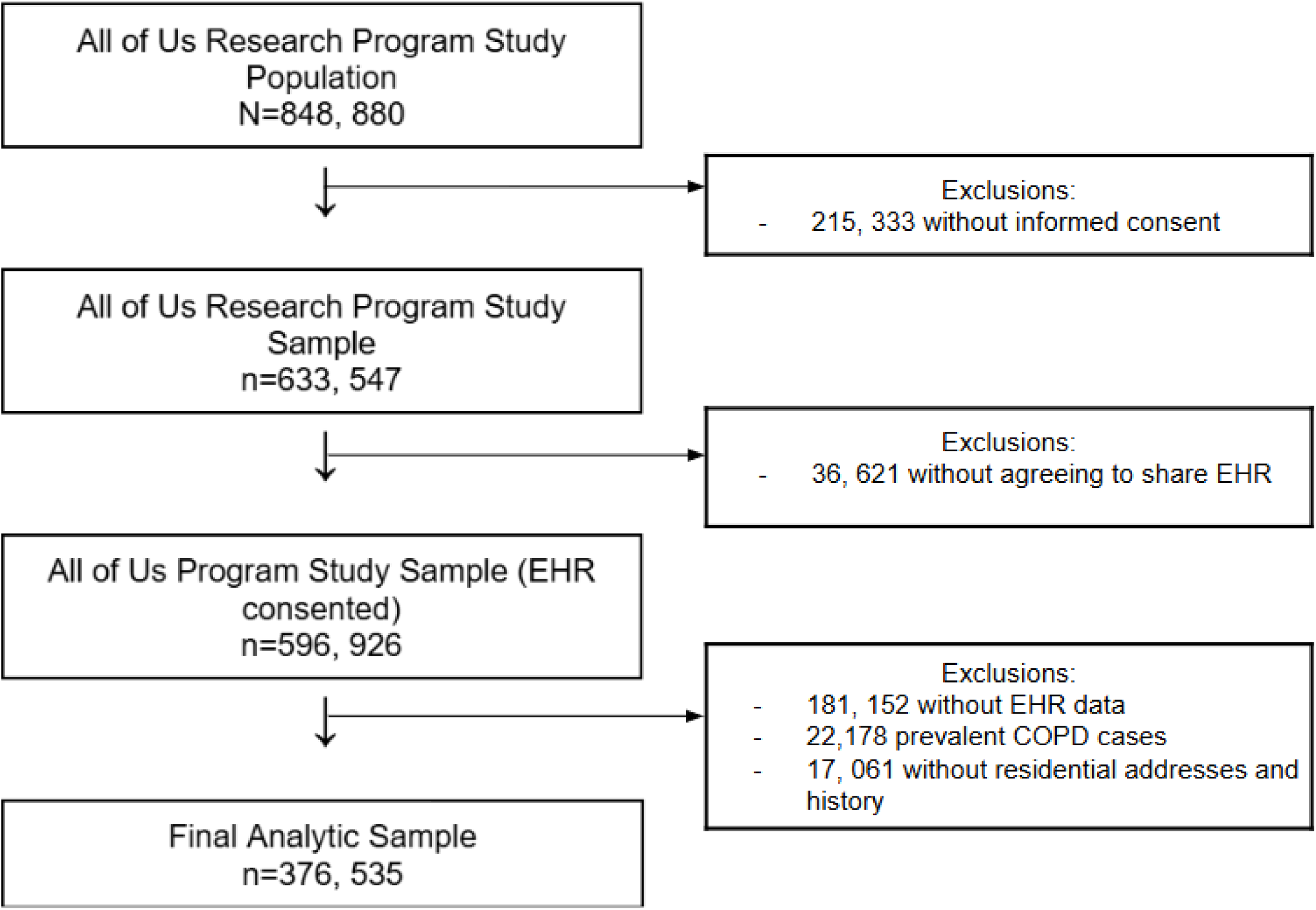
Inclusion for the study population and analytical cohort.

### Exposure Assessment and Data Linkage

Residential ambient air monitoring data measured using orbital satellites were obtained from the National Aeronautics and Space Administration (NASA)’s Socioeconomic Data and Applications Center (SEDAC) and Earth Observing System Data and Information System (EOSDIS). We extracted high-resolution, well-validated predictions generated using ensemble models for daily and annual concentrations of PM_2.5_ (µg/m^3^), ozone (ppb), and NO_2_ (ppb) at the zip code level across the contiguous U.S.^18^ ^19^. Using annual zip code-level PM_2.5_, ozone, and NO_2_ concentrations for the contiguous U.S. from 2000–2016, we calculated the concentrations of each pollutant for each three-digit postal region, by taking the mean of the concentrations of the encompassed five-digit zip codes.

Ozone and PM levels increase as temperatures rise due to increased gas-phase reaction rates ^20^ ^21^. As such, we extracted minimum, maximum, and average annual mean temperature from National Oceanic and Atmospheric Administration - National Centers for Environmental Information databases. These temperature data were linked to the participants’ residential zip code. Coordinate data for each zip code were extracted from Census 2010 data using the Python package uszipcode 1.0.15-8. The geometric center of each three-digit postal region was calculated from five-digit zip code data using the Haversine formula. The annual temperature values from the five closest satellites to the geometric center of each three-digit postal region were inverse distance weighted to estimate temperature metrics for each individual.

The environmental data values were assigned to each participant based on the duration of their residence at the reported address, as indicated by the enrollment date. Only mean annual air monitoring and temperature data corresponding to the years of residence at the reported address were included. Participants without available data for these linkage methods were assigned a missing value and excluded from the analyses.

### Spearman’s Correlation

To inform the associational and interaction analyses, correlation among independent variables was tested using Spearman’s correlation (r_s_). We observed slight inverse correlations between ozone and PM_2.5_ (r_s_=-0.29), ozone and NO_2_ (r_s_=-0.31), and annual average temperature and PM_2.5_ (r_s_=-0.07). Additionally, we found modest positive correlations between PM_2.5_ and NO_2_ (r_s_ =0.41), NO_2_ and annual average temperature (r_s_ = 0.18), and between ozone and annual average temperature (r_s_ =0.47).

### Outcome Assessment

The primary outcome of interest was incident COPD, which was ascertained using linked EHR data using validated International Classification of Diseases, Tenth Revision (ICD-10) code: J44 and Systematized Nomenclature of Medicine—Clinical Terms (SNOMED CT) code: 13645005. We defined incident COPD as the first recorded diagnosis. This approach was consistent with validated measurements in epidemiological studies, ensuring a comprehensive assessment of disease status ^22^.

### Statistical Analysis

#### Descriptive analyses

Descriptive statistics were used to summarize participant characteristics. Continuous variables were reported as means with standard deviations, and differences among groups were assessed using t-tests and ANOVA tests. Categorical variables were expressed as frequencies and percentages, and differences among groups were assessed using chi-squared tests. Heat maps were utilized to visualize the spatial distribution of participant enrollment, ozone levels, and incidence rates of COPD across the U.S. First, using individual-level data, we calculated the crude incidence rates over the follow-up period based on COPD diagnoses ascertained from EHR data. Subsequently, the 2000 U.S. standard population weights from the Census P25-1130 were used to estimate age- and sex-adjusted incidence rates. Data were aggregated at the state level, and values were normalized to facilitate comparisons. Heat maps were generated using R (ggplot2, tmap), employing a color gradient to represent varying intensities of distribution.

#### Exposure-disease association analyses

We used multivariable Cox proportional hazards models to estimate the hazard ratios (HRs) and 95% confidence intervals (CIs) of incident COPD in relation to ozone concentrations, categorized as quartiles as well as continuous (per ppb) to assess linear trends. Time-on-study (years) was used as the time scale. We adjusted for age at enrollment (years), sex (male, female), race (White, Black or African American, Asian, Other), Hispanic status, income (<$75,000, $75,000–$150,000, >$150,000), education level (high school graduate or less, some college, college graduate, or advanced degree or higher), BMI (kg/m^2^), smoking status (never- or ever-smoker), smoking pack-years (py), and PM_2.5_ concentrations (µg/m^3^). Additionally, we conducted stratified analyses to evaluate heterogeneity among demographic subgroups defined by age, sex, race, smoking status, and annual average household income as a proxy for SES. These subgroup analyses were specified *a priori*. Subsequently, we fitted multiplicative interaction terms between ozone, demographic characteristics, PM_2.5_, and NO_2_ in the Cox models to provide additional evidence for heterogeneity or potential effect modification. We visually evaluated violations of proportional hazards assumptions by inspecting the Schoenfeld residuals over the follow-up time and did not find evidence of violations for the main effect of ozone. Two-sided P-values <0.05 were considered statistically significant.

We conducted exploratory analyses, using multivariable Cox proportional hazards models, for the joint effect between ozone, NO_2_ concentrations (ppb), and temperature on COPD risk. Ozone concentrations were first dichotomized at the median into low and high exposure groups (≤ and > median ppb), while annual average temperature was dichotomized into low and high temperature groups (≤ and >median °C). These categories were then combined into a 4-category cross-classification variable: low ozone/low temperature (reference group), high ozone/low temperature, low ozone/high temperature, and high ozone/high temperature.

#### Accounting for Potential Volunteer Selection Bias

The AoU research program is not a population-based study with probabilistic sampling. Rather, AoU enrolled volunteers, which helps maximize the potential to recruit a large number of participants needed to investigate complex interactions in the omics era ^16^ ^17^ ^22^. To account for potential volunteer bias in participant enrollment, cell weighting was applied to each participant in the statistical models by calibrating the marginal distribution of various demographic factors using the 2018-2022 American Community Survey (ACS) Public Use Microdata Sample (PUMS) as a reference. The weight flooring and ceiling were set at the 5th and 95th percentiles. The first set of weights adjusted for age, race, Hispanic status, education, and income at the state level. The second set of weights contained the variables from the first set and additionally adjusted for marital status, military status, insurance status, and immigrant status.

## Results

### Baseline Characteristics

The distribution of key baseline characteristics differed by COPD status (Table 1). On average, incident COPD cases were older (60.9 years vs. 51.8 years), had a larger proportion of males (44.5% vs.37.7%), and had a larger proportion of Black participants (27.5% vs. 18.0%) compared to the non-cases. Further, COPD cases had lower educational attainment (41.5% vs. 26.8% with high school education or less) and reported lower annual income (54.8% vs. 38.0% earning below $75,000 per year). Additionally, COPD cases had higher BMI compared to those without COPD (30.8 kg/m^2^ vs. 29.9 kg/m^2^). Never-smokers made up only 26.7% of incident COPD cases compared to 58.5% among non-cases. COPD cases had a higher proportion of ever-smokers (70.2% vs. 38.1%) and reported higher average pack-years (py) of smoking (18.9 py) compared to non-cases (5.20 py).

**Table 1:** Baseline characteristics of incident COPD cases and non-cases in the All of Us research program.

| **Characteristic** | **Overall (N=376535)** | **Incident COPD Cases (n=7907)** | **Non-cases (n=368628)** |
| --- | --- | --- | --- |
| Age, years, mean (SD) | 52.0 (17.0) | 60.9 (12.3) | 51.8 (17.1)* |
| Age at diagnosis of COPD, years, mean (SD) | 62.6 (12.4) | 62.6 (12.4) | — |
| Sex |  |  |  |
| Male, n (%) | 142377 (37.8) | 3518 (44.5) | 138859 (37.7)* |
| Female, n (%) | 227454 (60.4) | 4249 (53.7) | 223205 (60.6) |
| Race |  |  |  |
| White, n (%) | 216153 (57.4) | 4416 (55.8) | 211737 (57.4)* |
| Black, n (%) | 68478 (18.2) | 2174 (27.5) | 66304 (18.0) |
| Asian, n (%) | 11620 (3.1) | 59 (0.7) | 11561 (3.1) |
| Hispanic Status |  |  |  |
| Hispanic or Latino, n (%) | 68351 (18.2) | 954 (12.1) | 67397 (18.3)* |
| Not Hispanic or Latino, n (%) | 297159 (78.9) | 6644 (84.0) | 290515 (78.8) |
| Smoking Status |  |  |  |
| Ever smoker, n (%) | 146107 (38.8) | 5552 (70.2) | 140555 (38.1)* |
| Never smoker, n (%) | 217652 (57.8) | 2114 (26.7) | 215538 (58.5) |
| Smoking pack-years, py, mean (SD) <sup>1</sup> | 5.4 (13.9) | 18.9 (25.4) | 5.2 (13.5)* |
| Body mass index, kg/m<sup>2</sup>, mean (SD) | 29.9 (7.6) | 30.8 (8.5) | 29.9 (7.6)* |
| Highest education level attained, n (%) |  |  |  |
| Highschool, GED, or below | 102048 (27.1) | 3284 (41.5) | 98764 (26.8)* |
| Undergraduate | 97198 (25.8) | 2384 (30.2) | 94814 (25.7) |
| College | 86645 (23.0) | 1106 (14.0) | 85539 (23.2) |
| Advanced degree | 81516 (21.6) | 868 (11.0) | 80648 (21.9) |
| Annual income, n (%) |  |  |  |
| Below \$75,000 | 144593 (38.4) | 4333 (54.8) | 140260 (38.0)* |
| \$75,000 to \$150,000 | 112752 (29.9) | 1436 (18.2) | 11316 (30.2) |
| Above \$150,000 | 44440 (11.8) | 268 (3.4) | 44172 (12.0) |
| Duration of Residence at Baseline, n (%) |  |  |  |
| 0 to 2 years | 121327 (32.23) | 2591 (32.8) | 118736 (32.2) |
| 3 to 5 years | 66864 (17.8) | 1410 (17.8) | 65454 (17.8) |
| 6 to 10 years | 51827 (13.8) | 1084 (13.7) | 50743 (13.8) |
| 11 to 20 years | 57179 (15.2) | 1114 (14.1) | 56065 (15.2) |
| >20 years | 73879 (19.6) | 1526 (19.3) | 72353 (19.6) |
| Annual average temperature, °C, mean (SD) | 14.4 (5.2) | 15.2 (5.4) | 14.4 (5.2)* |
| Annual fine particulate matter 2.5 (PM<sub>2.5</sub>),<br>µg/m<sup>3</sup>, mean (SD) | 10.6 (2.4) | 10.4 (2.6) | 10.6 (2.4)* |
| Annual nitrogen dioxide (NO<sub>2</sub>), ppb, mean<br>(SD) | 25.8 (9.8) | 27.3 (9.9) | 25.8 (9.8)* |
| Annual ground-level ozone, ppb, mean (SD) | 38.7 (4.3) | 39.3 (4.8) | 38.7 (4.3)* |

<sup>1</sup>Based on an overall sample size of 321, 025 participants. \*p<0.001.

Residential duration at baseline was similar between COPD cases and the overall study population (19.3% vs 19.6% living more than 20 years at their residence at enrollment). COPD cases were exposed to higher ambient ozone concentrations at their residential location (39.3 ppb vs. 38.7 ppb) and had higher mean annual average temperatures (15.2°C vs. 14.4°C) compared to the overall study population. Additionally, NO_2_ levels were slightly higher among COPD cases compared to the overall study population (27.3 ppb vs. 25.8 ppb). Annual PM_2.5_ concentrations were similar by COPD status.

### Regional Patterns in Ozone Concentrations and Incidence Rates of COPD

Ozone concentrations were higher in Arizona in the western regions as well as Tennessee in the eastern regions (Figure 2A). This approximately corresponded with the higher age- and sex-adjusted incidence rates in these states (Figure 2D).

**Figure 2:**
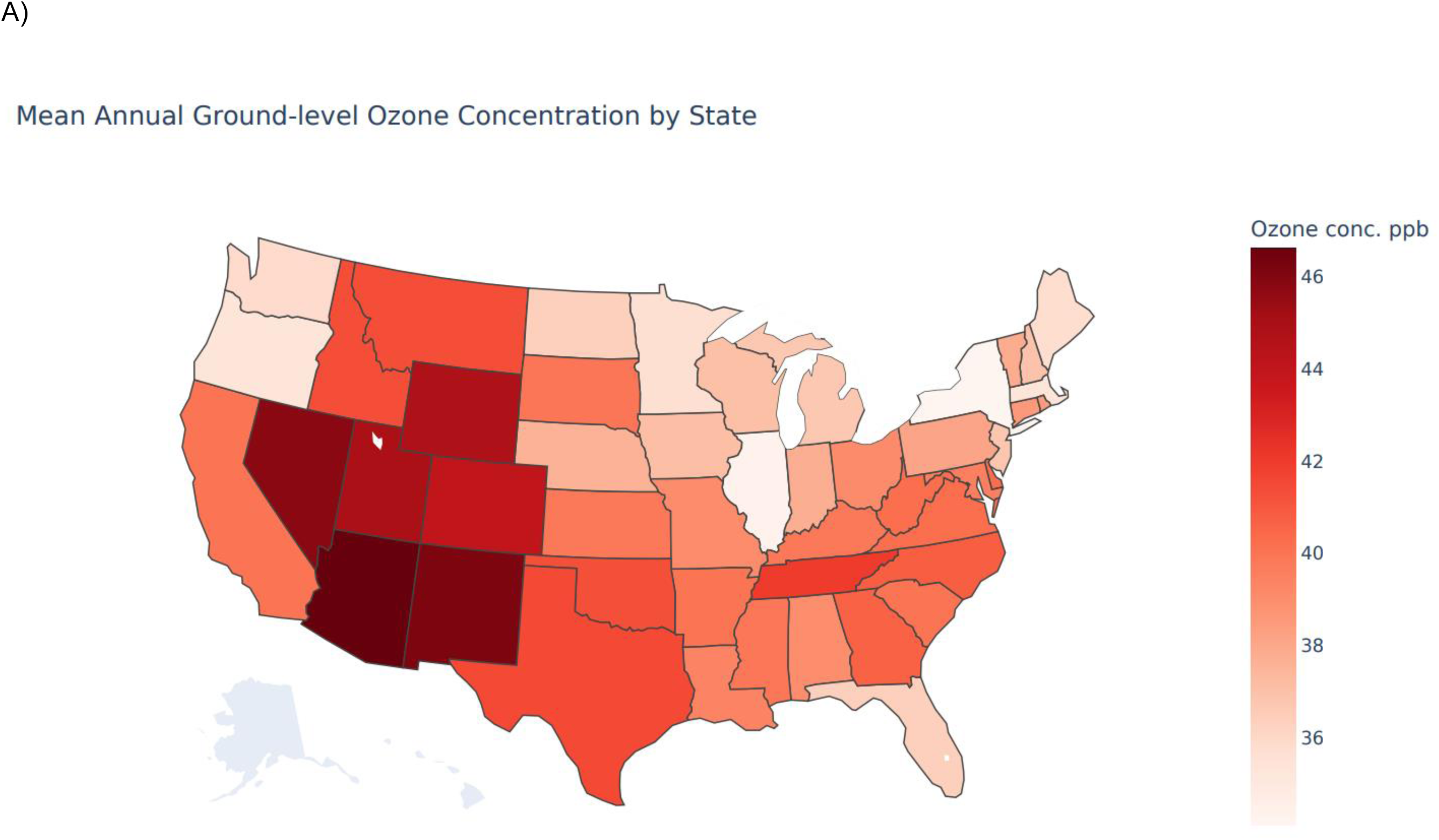
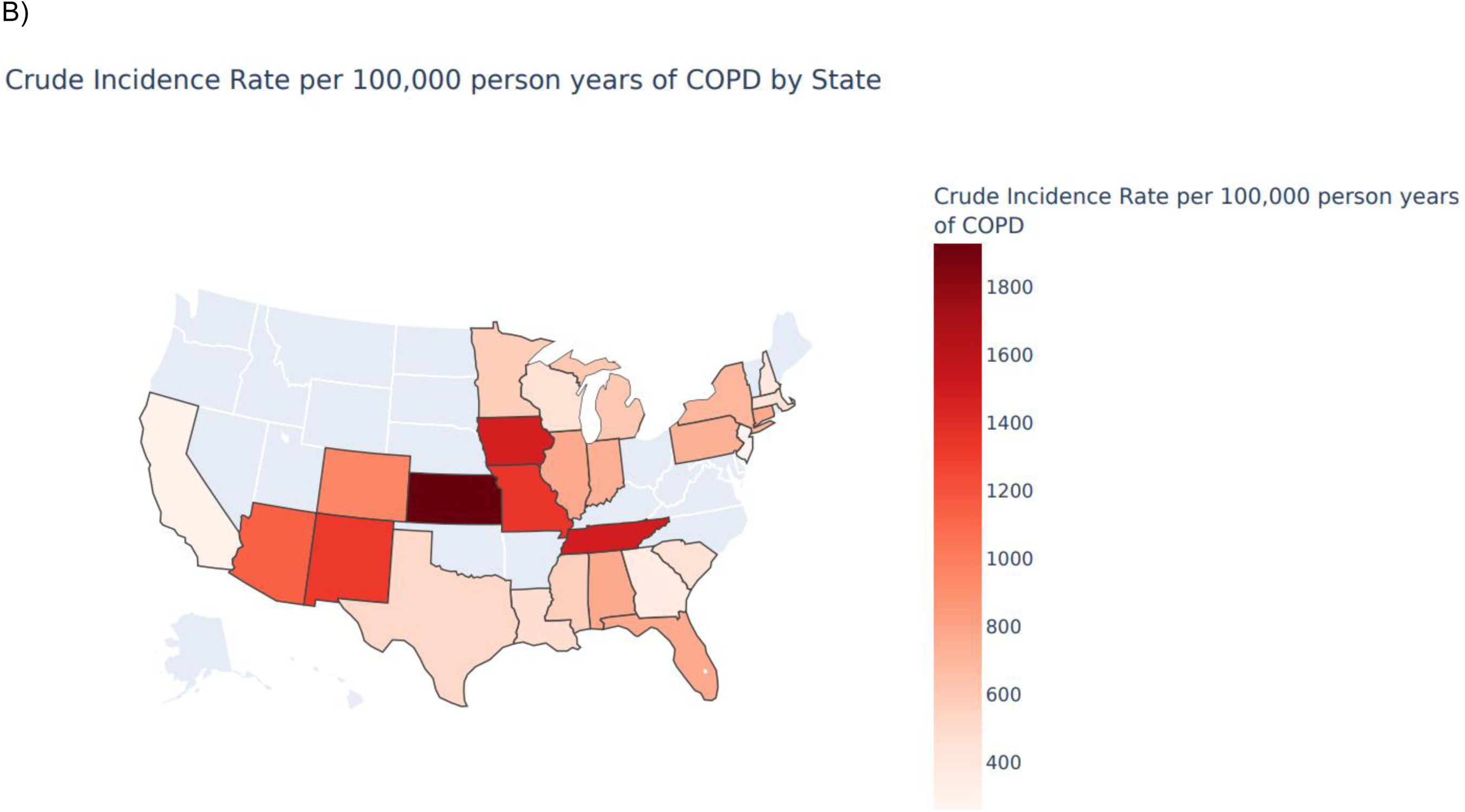
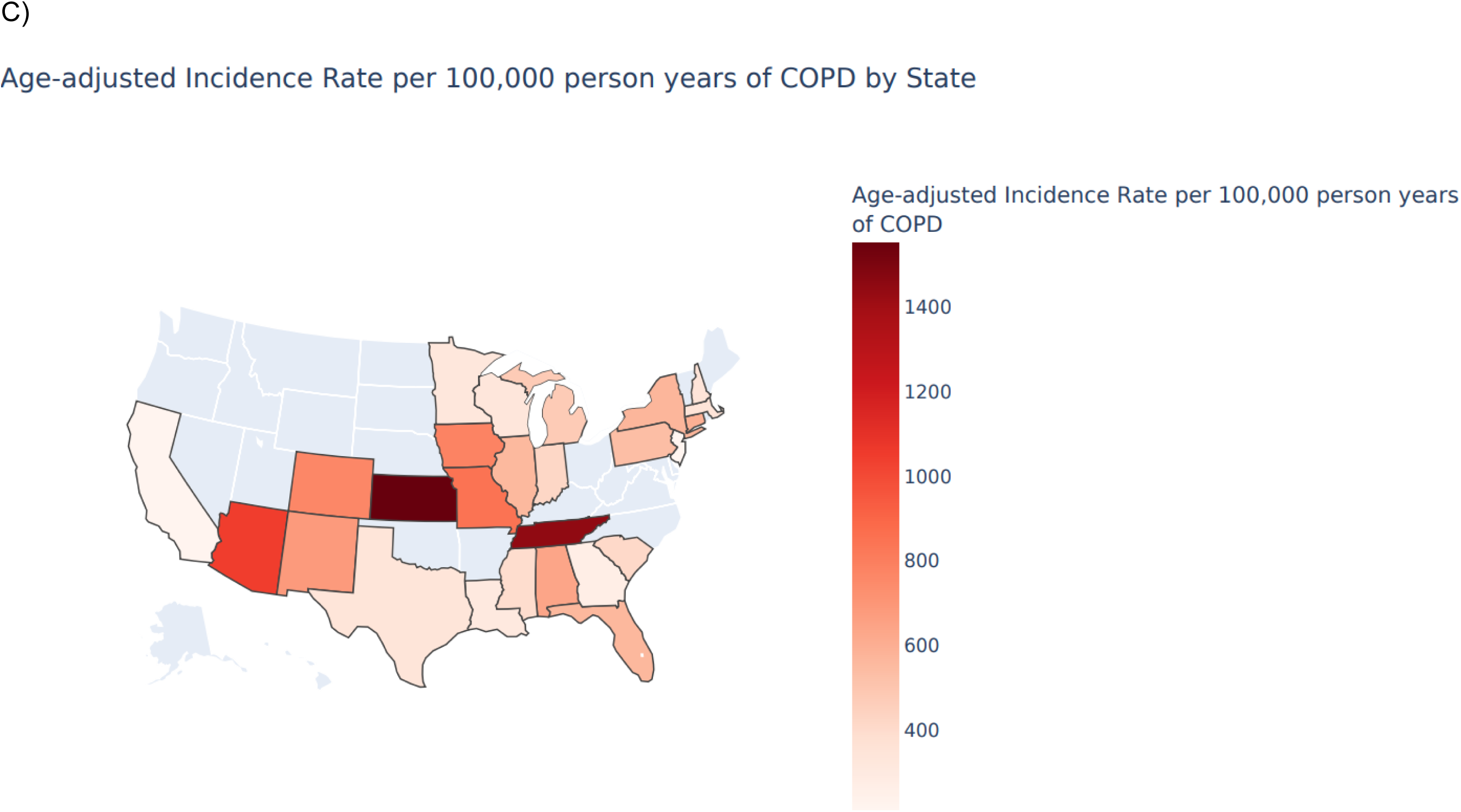
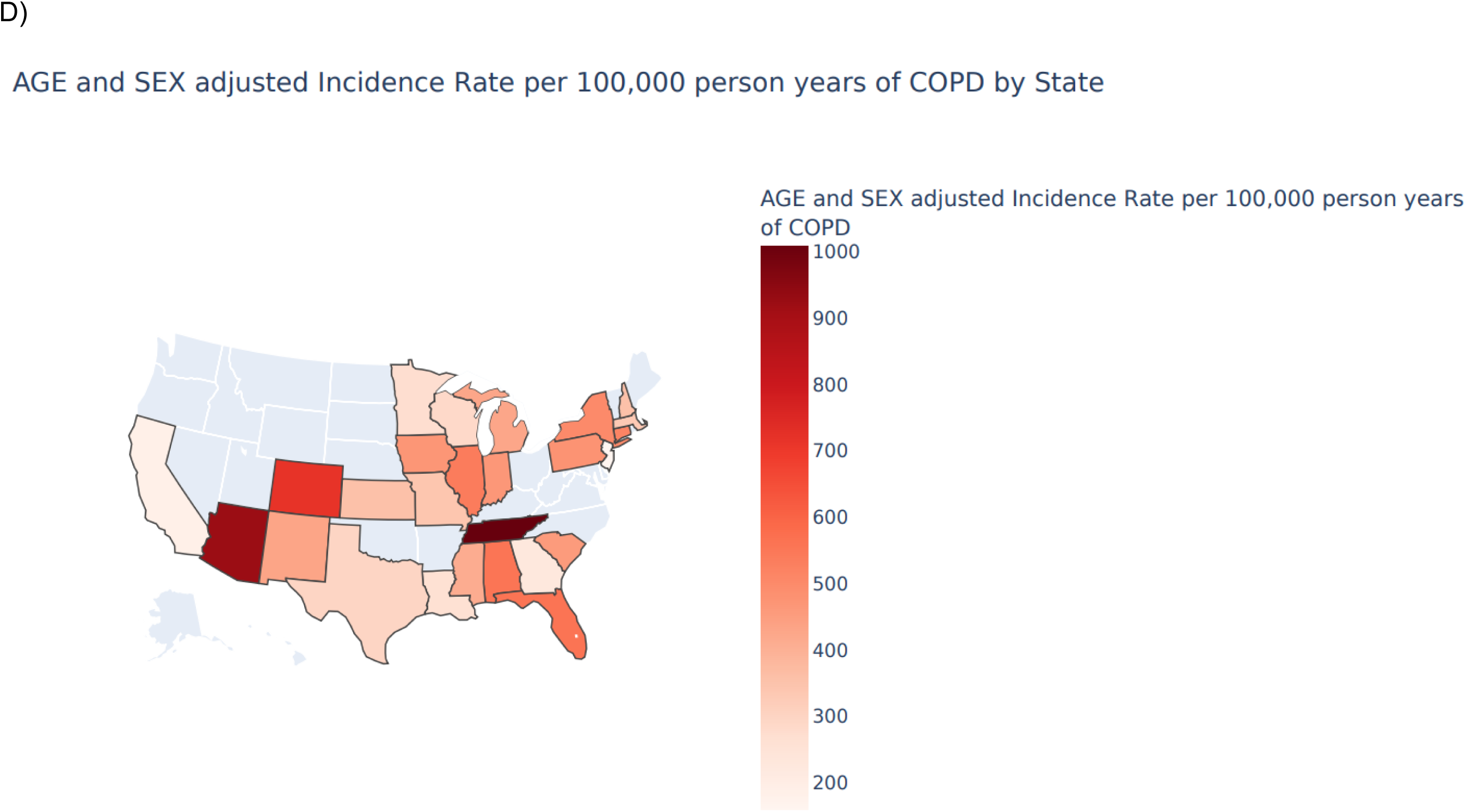
Distribution of A) ozone concentration between 2000—2016, B) crude COPD incidence rate, C) age-adjusted COPD incidence rate, and D) age- and sex-adjusted COPD incidence rate by state in the conterminous United States.

### Overall Association between Residential Ozone Exposure and Risk of COPD

We identified 7,907 incident COPD cases over an average 4-year follow-up. The mean age at diagnosis of COPD was 62.6 years. We observed a positive, non-monotonic relationship between residential ozone and COPD risk (Table 2). The association was most apparent in the third (HR=1.02, 95% CI: 0.95, 1.11) and highest quartile of ozone (HR: 1.09, 95% CI: 1.01, 1.18).

**Table 2:**
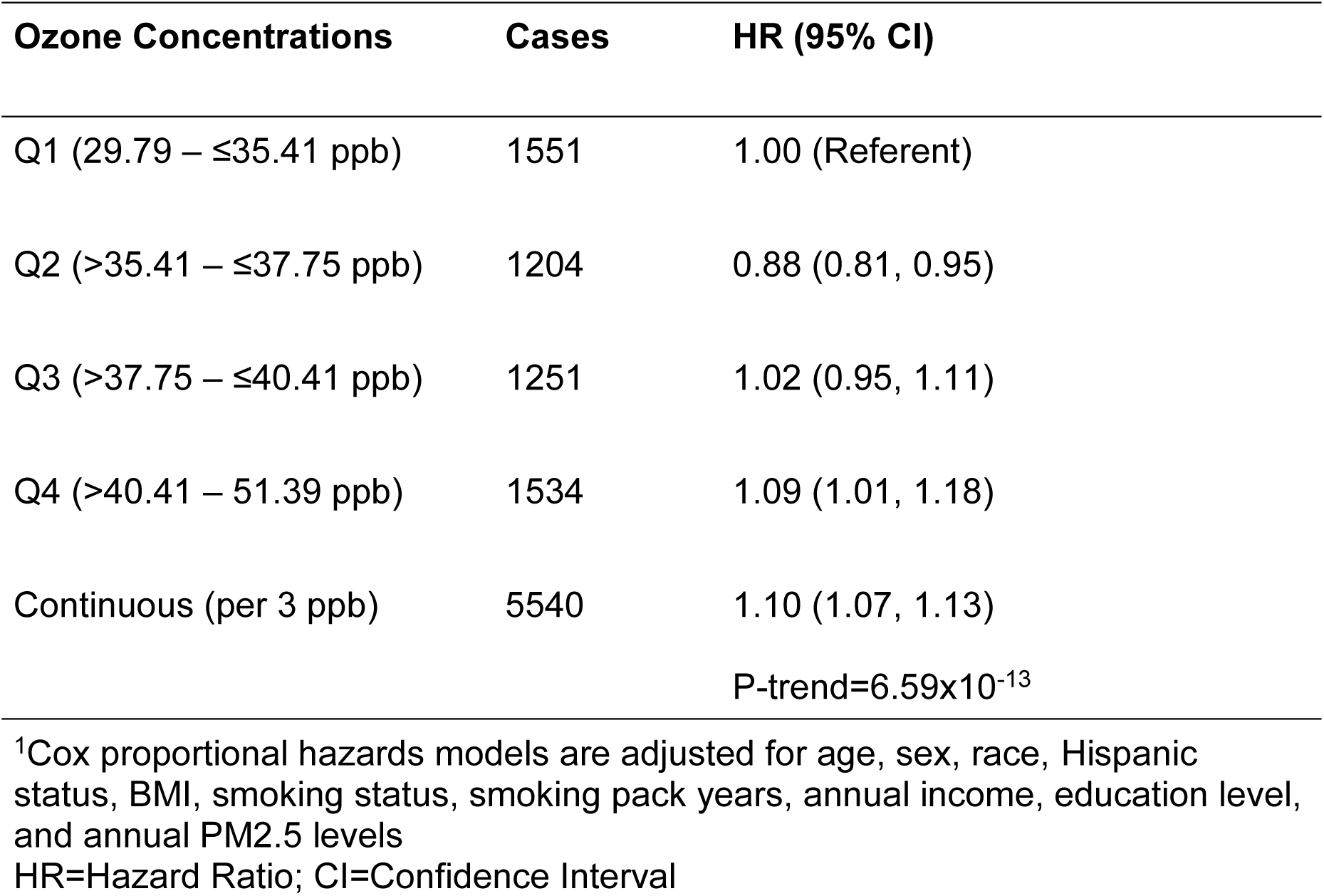
Overall associations between residential ground-level ozone concentrations and incident COPD risk in the All of Us research program^1^.

### Heterogeneity in Ozone-COPD Associations by Demographic Subgroups

#### Sex and Smoking Status

We found notable differences in the exposure-response relationship between ozone and COPD across subgroups defined by sex and smoking status (Table 3; P-interaction_Males: Ozone x Smoking_=0.01; P-interaction_Females: Ozone x Smoking_=0.06). The ozone-COPD trend was most pronounced and monotonic among male never-smokers ((HR_per 3 ppb_=1.14, 95% CI: 1.08, 1.21) P-trend_continuous_=6.62×10^-6^), with the highest quartile of ozone exposure being significantly associated with COPD risk (HR=1.39, 95% CI: 1.11−1.74). Among male ever smokers, the ozone-COPD trend was non-monotonic, with the highest quartile of ozone exposure being significant (HR=1.14, 95% CI: 1.01, 1.29). Among females, there was some evidence of a U-shaped relationship among female never-smokers, with a nadir at the second quartile of ozone (HR=0.73, 95% CI: 0.61, 0.87) (Table 3).

**Table 3:**
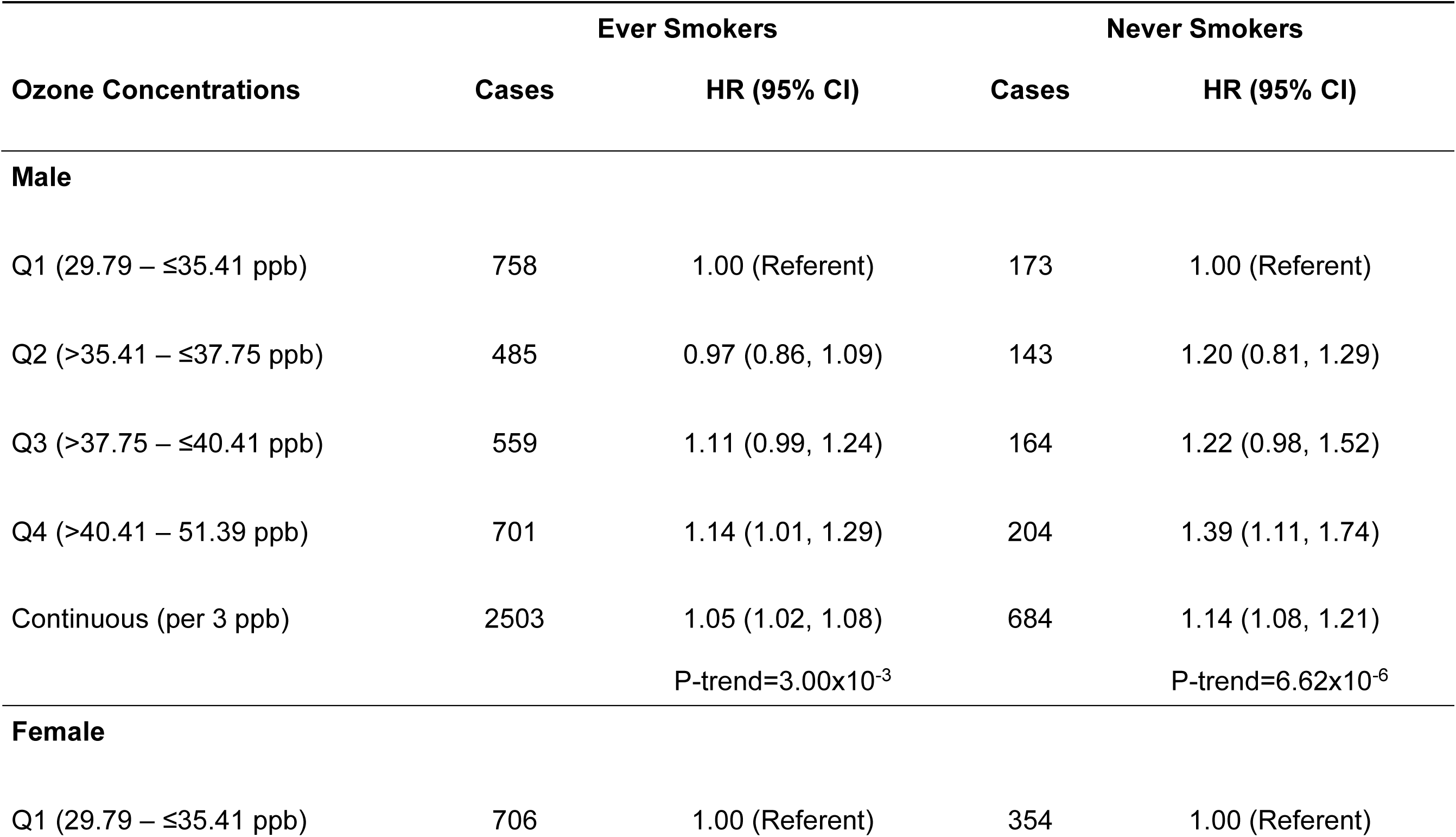
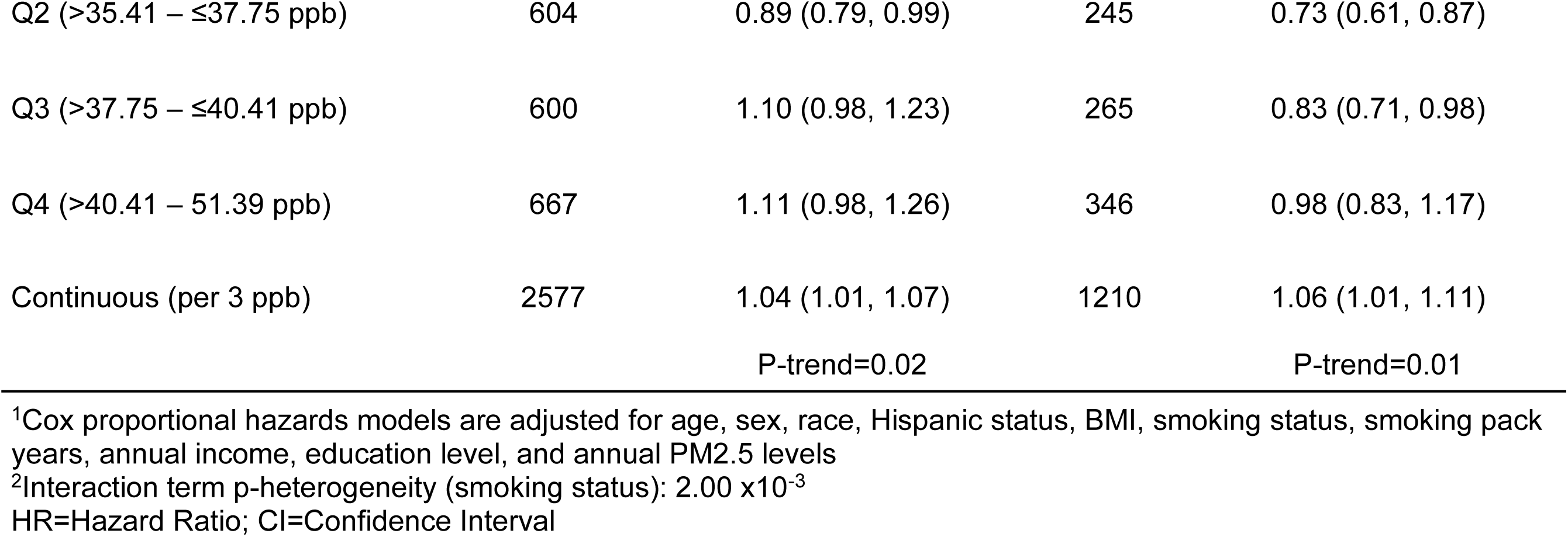
Associations between residential ground-level ozone concentrations and incident COPD risk by sex and smoking status in the All of Us research program^1, 2^.

#### Race and Sex

The observed ozone-COPD associations were similar qualitatively across subgroups defined by sex and race, with some evidence for heterogeneity (Table 4). Among males, White participants ((HR_per 3 ppb_=1.09, 95% CI:1.05, 1.13) P-trend_continuous_=2.76×10^-5^) had similar but more pronounced trends compared with Black or African American individuals ((HR_per 3 ppb_=1.04, 95%CI: 0.98, 1.12) P-trend_continuous_=0.22; P-interaction=0.13). Among females, the ozone-COPD associations at the highest quartile were more apparent among White individuals (HR=1.28, 95% CI: 1.08, 1.52).

**Table 4:**
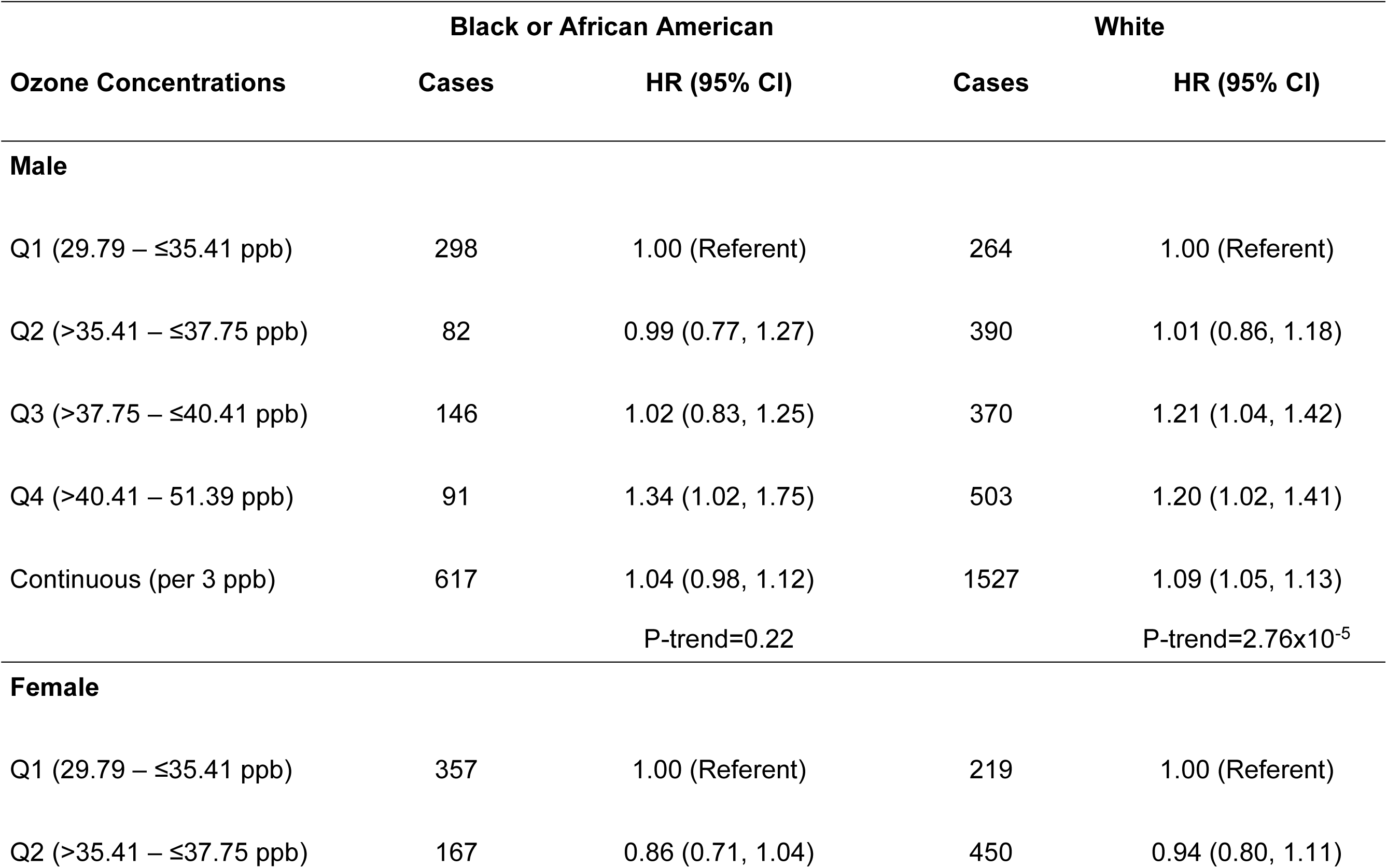
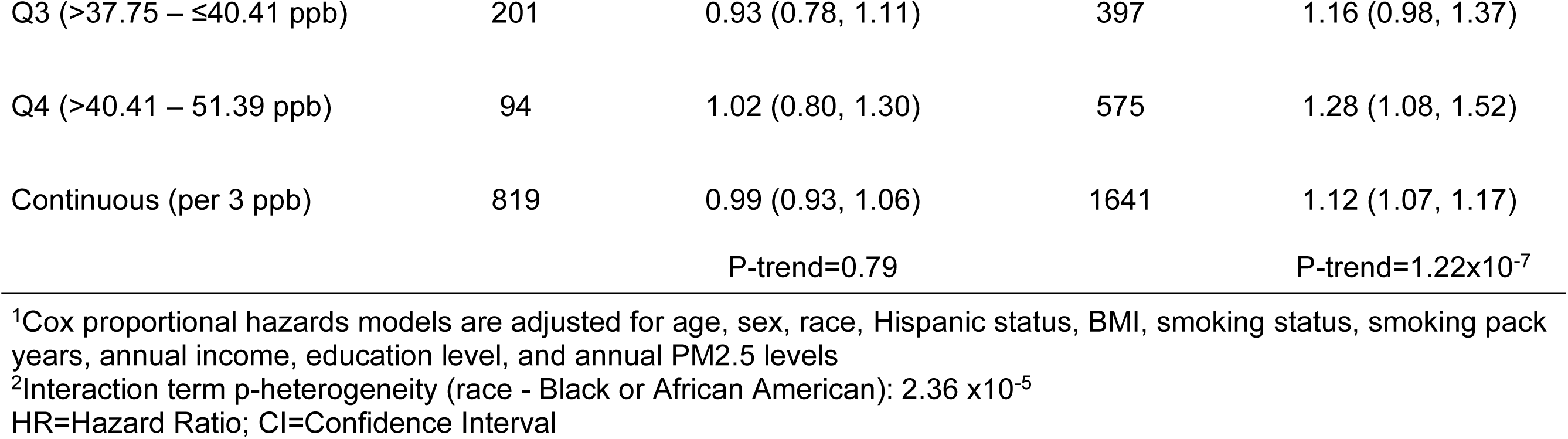
Associations between residential ground-level ozone concentrations and incident COPD by race and sex in the All of Us research program^1, 2^.

#### Age and Sex

We found that the ozone-COPD association was more apparent among male subgroups <65 years of age (Table 5); however, the tests for interaction were non-significant (P-interaction_Males: Ozone x Age_=0.08; P-interaction_Females: Ozone x Age_=0.80). Among younger males (20 to 44 years), we observed a positive but non-monotonic relationship. Here, the third quartile of ozone was associated with increased COPD risk (HR=1.56, 95% CI: 1.00, 2.41), while the association in the highest quartile was more suggestive (HR=1.50, 95% CI: 0.96, 2.35). Among middle-aged males (45 to 65 years), we observed a positive monotonic exposure-response relationship ((HR_per 3 ppb_=1.08, 95% CI: 1.03, 1.13) P-trend_continuous_=0.001), with the strongest association evident in the fourth quartile (HR=1.24, 95% CI: 1.04, 1.47).

**Table 5:**
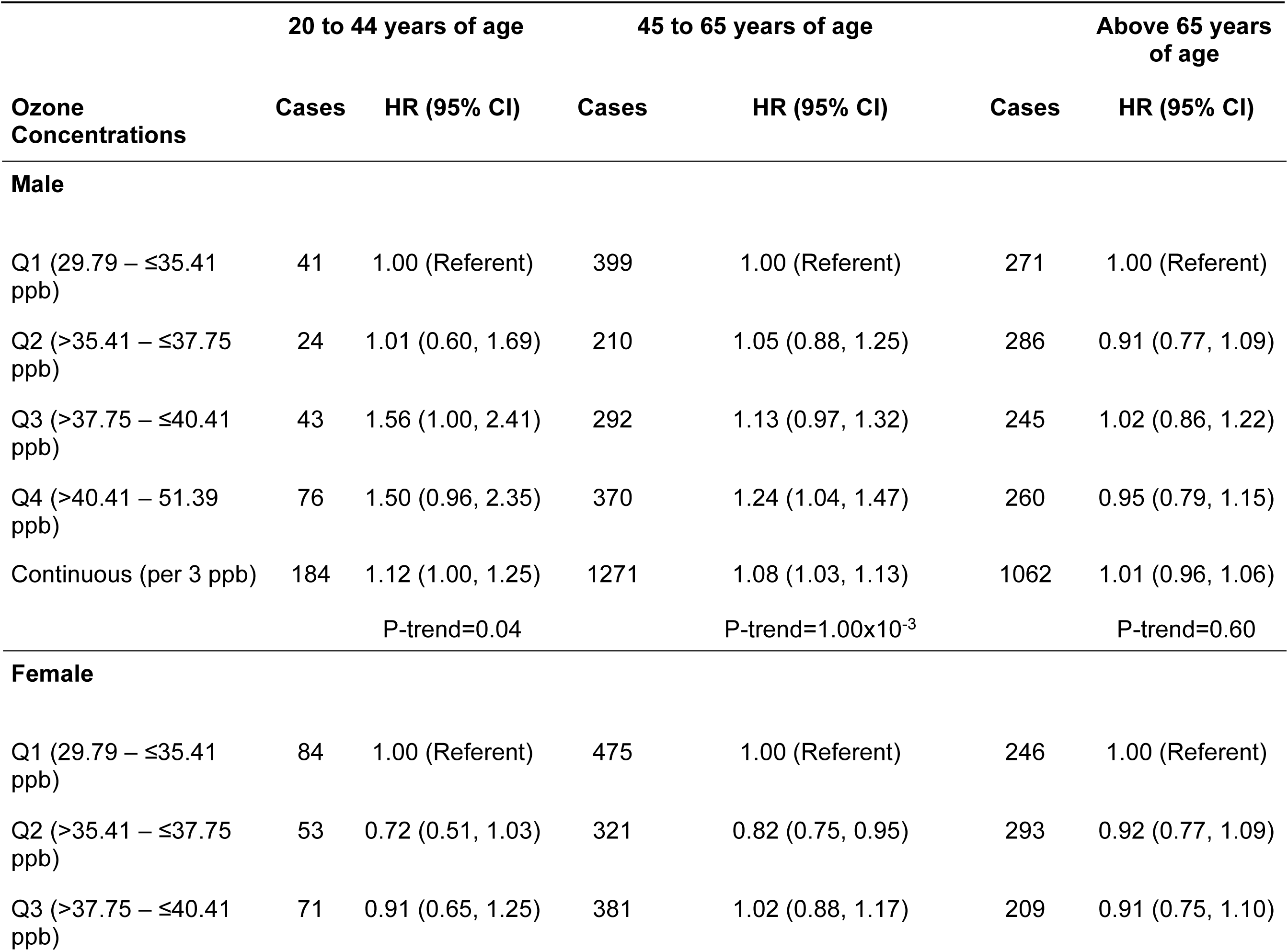
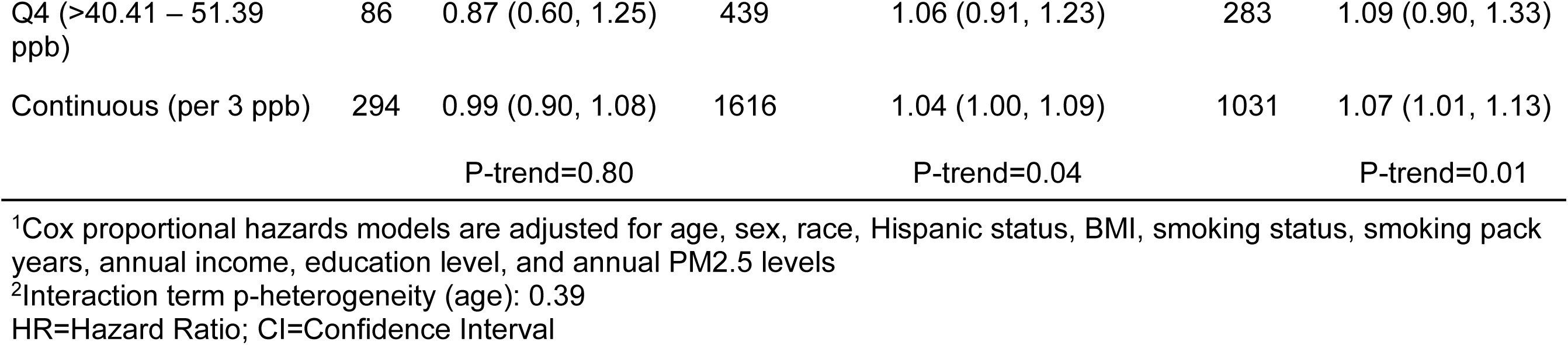
Associations between residential ground-level ozone concentrations and incident COPD risk by age and sex group in the All of Us research program^1, 2^.

#### Race and Income

We found that the ozone-COPD association was more apparent among certain subgroups defined by socioeconomic status and race (Supplementary Table 1); however, the tests for interaction did not show evidence of statistical heterogeneity (P-interaction_White: Ozone x Middle Income_=0.20; P-interaction_White: Ozone x Upper Income_=0.09; P-interaction_Black: Ozone x Middle Income_=0.95; P-interaction_Black: Ozone x Upper Income_=0.31). Among White participants, we observed positive non-monotonic trends among those in the lowest income group, with the third (HR=1.26, 95% CI: 1.06, 1.50) and fourth (HR=1.32, 95% CI: 1.11, 1.57) quartile of ozone being significant. The middle-income group showed similar trends, though slightly attenuated.

#### Sex and Income

We found that the ozone-COPD associations were more apparent among certain subgroups defined by sex and annual income (Supplementary Table 2); however, we found no evidence of statistical interaction (P-interaction_Males: Ozone x Middle Income_=0.30; P-interaction_Males: Ozone x Upper Income_=0.50; P-interaction_Females: Ozone x Middle Income_=0.06; P-interaction_Females: Ozone x Upper Income_=0.20). Among males with lower annual income, we found a positive non-monotonic relationship, with the highest quartile of ozone being significantly associated with COPD risk (HR=1.17, 95% CI: 1.00, 1.39). A similar but weaker association was observed among middle-income males. We did not detect ozone-COPD associations among higher-income males; however, the case numbers were limited. Among females, we detected a positive, non-monotonic relationship among those of middle-income.

### Ozone-COPD Association Accounting for Atmospheric Conditions

We observed a weak negative interaction between ozone and NO_2_ (β =-0.01 (P-interaction=1.06×10^-11^)). Additionally, we observed a modest negative interaction between ozone and PM_2.5_ (β=-0.04 (P-interaction=2.00×10^-16^)), and a positive interaction between ozone and temperature (β=0.01 (P-interaction=3.53×10^-8^)).

Accounting for annual average temperature and NO_*2*_ did not diminish the observed ozone-COPD associations (Supplementary Table 3). However, we did not find compelling evidence that ozone, temperature, and NO_2_ acted synergistically in cross-classification analyses. When compared to individuals exposed to low ozone and low temperatures, those with high ozone exposure and high temperatures had increased COPD risk (HR=1.21, 95% CI: 1.11, 1.30). Similar COPDs risks were observed for those with low ozone exposure and high temperatures (HR=1.31, 95% CI: 1.18, 1.45) and high ozone and low temperatures (HR=1.20, 95% CI: 1.06, 1.35).

### Weighting to Account for Potential Volunteer Selection Bias

The observed ozone-COPD associations were robust even after applying different weighting techniques to correct for potential volunteer selection bias (Supplementary Table 4). Across all weighting methods, the non-monotonic relationship remained significant. Quartile analyses showed nominally higher COPD risk in both weighting methods ((HR_Q4_=1.16, 95% CI: 1.03, 1.31; HR_Q4_=1.19, 95% CI: 1.03, 1.37)).

## Discussion

In the prospective AoU research program, we found that higher residential ozone concentrations were associated with an increased risk of incident COPD, even at exposure levels consistently below the current U.S. regulatory limit. Our findings remained robust after additional adjustment for co-exposures and atmospheric conditions, as well as correcting for potential volunteer bias. We found some evidence that NO_2_ and PM_2.5_ slightly attenuated the ozone-COPD association, whereas temperature strengthened the association. However, further analyses did not support a synergistic relationship between ozone and temperature. By integrating additional environmental variables, our study provided a more nuanced and unbiased understanding of the intersection between environmental factors and COPD risk.

When examining demographic subgroups, we found evidence suggesting that the respiratory effects of ozone might be more apparent among males below 65 years of age, males of low-middle income, and especially male never-smokers. Environmental and occupational epidemiology studies have shown that the respiratory effect of smoking is so strong, that it can mask the contribution of environmental exposures ^23^. As such, investigating never-smokers allows for a more unbiased estimate of the ozone-COPD relationship ^24^ ^25^. Building on recent research on non-smoking risk factors in the development of COPD ^26^, our study provides important new insights into the respiratory effects of ozone that support the need for greater focus on COPD risk in never-smokers ^26^.

Among male never-smokers, we observed a striking monotonic exposure-response relationship between ozone and COPD risk. However, the association was unclear among female never-smokers. These differences may be due to varying lifestyle, occupational, and other environmental exposures ^27 28^. Although COPD prevalence is typically higher among female never-smokers, studying male never-smokers is important because research suggests they may experience greater disease severity, with earlier onset and more pronounced respiratory symptoms ^29 30^. Furthermore, there is a notable research gap on COPD among male never-smokers, highlighting the need for comprehensive understanding of the disease across sexes ^31 32^.

We posit that the pathobiological mechanisms underlying the association between ozone exposure and COPD were primarily related to ozone-induced oxidative stress, airway inflammation, and epithelial damage. Previous studies have shown that ozone-induced oxidative stress and inflammation have been shown to play pivotal roles in disease initiation, progression, and exacerbations of COPD ^33 34 35^. More specifically, ozone exposure may disrupt lung homeostasis by generating reactive oxygen species (ROS), which led to inflammatory cascades and structural lung impairment ^36 37^.

This study had several notable strengths. First, the AoU research program is a data-rich mega cohort of U.S. adults with linked EHRs and questionnaire data. The purposeful recruitment of minority populations from across the country allowed for the analysis of subgroup differences with greater granularity and statistical power. Second, our integration of advanced satellite-based air monitoring and atmospheric data improved the accuracy and resolution of ozone measurements, other chemical compounds, and temperature assessments over extended periods, enhancing the reliability of our findings.

Our study had some limitations. First, residual and unmeasured confounding may be present. However, we performed rigorous adjustments for potential confounders based on prior studies and conducted stratification, attenuation, and sensitivity analyses to assess the influence of residual confounding from smoking, co-exposures to NO_2_ and PM_2.5_, and other factors ^38^. Adjusting for NO_2_, a correlated surrogate for other chemical compounds from traffic and industrial sources, helped indirectly adjust for the cumulative impact of air pollutants ^33^. Further, our main findings were consistent with previous studies, which increases our confidence in the results ^30 39 40^. Second, geolocation of residence was provided only as a three-digit postal region to protect participant privacy. However, ozone generally has lower spatial variation compared to other pollutants such as PM_2.5_ and NO_2_ ^41 42 43^. Use of the 3-digit postal region along with a participant’s residential history might have led to some exposure misclassification from masking the spatial heterogeneity of ozone. However, this misclassification would be non-differential and likely lead to conservative underestimates of the true effect. Due to data availability, we were only able to assign exposure values based on a limited duration of residence. We acknowledge that this may introduce time-trend bias, as ambient ozone levels have generally declined over the study period; as such, this could result in lower average exposures among younger or more mobile participants. Third, ozone might not have been measured in the appropriate etiologic window relevant to COPD onset, as chronic respiratory diseases often develop over extended periods due to cumulative exposures. Ozone estimates were available only for the residential locations reported at the time of enrollment and we had to assume that these levels were consistent with participants’ residences in prior years. Given that the study population was residentially stable, with nearly two-thirds of the participants living at the current address for ≥3 years and almost 20% residing there for >20 years, we are confident that ozone concentrations were captured in etiologic windows relevant to COPD onset. To minimize the potential bias of this concern, we utilized long-term air pollution exposure estimates, incorporating annual averages over multiple years to better capture the cumulative exposure burden relevant to COPD pathogenesis. Additionally, ascertainment bias may be a concern, as individuals with higher healthcare utilization may be more likely to receive a COPD diagnosis, potentially skewing incidence estimates. However, given the generally uniform access to care in our study population, we expect this bias to be limited; future studies could further mitigate this issue by incorporating standardized screening protocols or adjusting for healthcare utilization metrics. Caution is warranted when interpreting these findings. Even though our study population was large, we cannot discount the possibility of statistical power issues for some demographic groups. Future studies in well-powered, population-based cohorts are needed to replicate these findings among granular demographic subgroups.

In conclusion, we found a robust relationship between residential ozone concentrations and COPD risk. The observed associations were more prominent among certain demographic subgroups, particularly male never-smokers. Our findings provide important insights into the relationship between ozone and newly-diagnosed COPD, highlighting that some demographic subgroups might be more impacted by certain air pollutants. Although ozone concentrations in our study remained consistently below the current U.S. regulatory limit, the observed increase in COPD risk supports the need to reassess air quality standards in the context of chronic respiratory health outcomes.

## Data Availability

All data produced in the present study are available upon reasonable request to the authors

---

**Supplementary Table 1:**
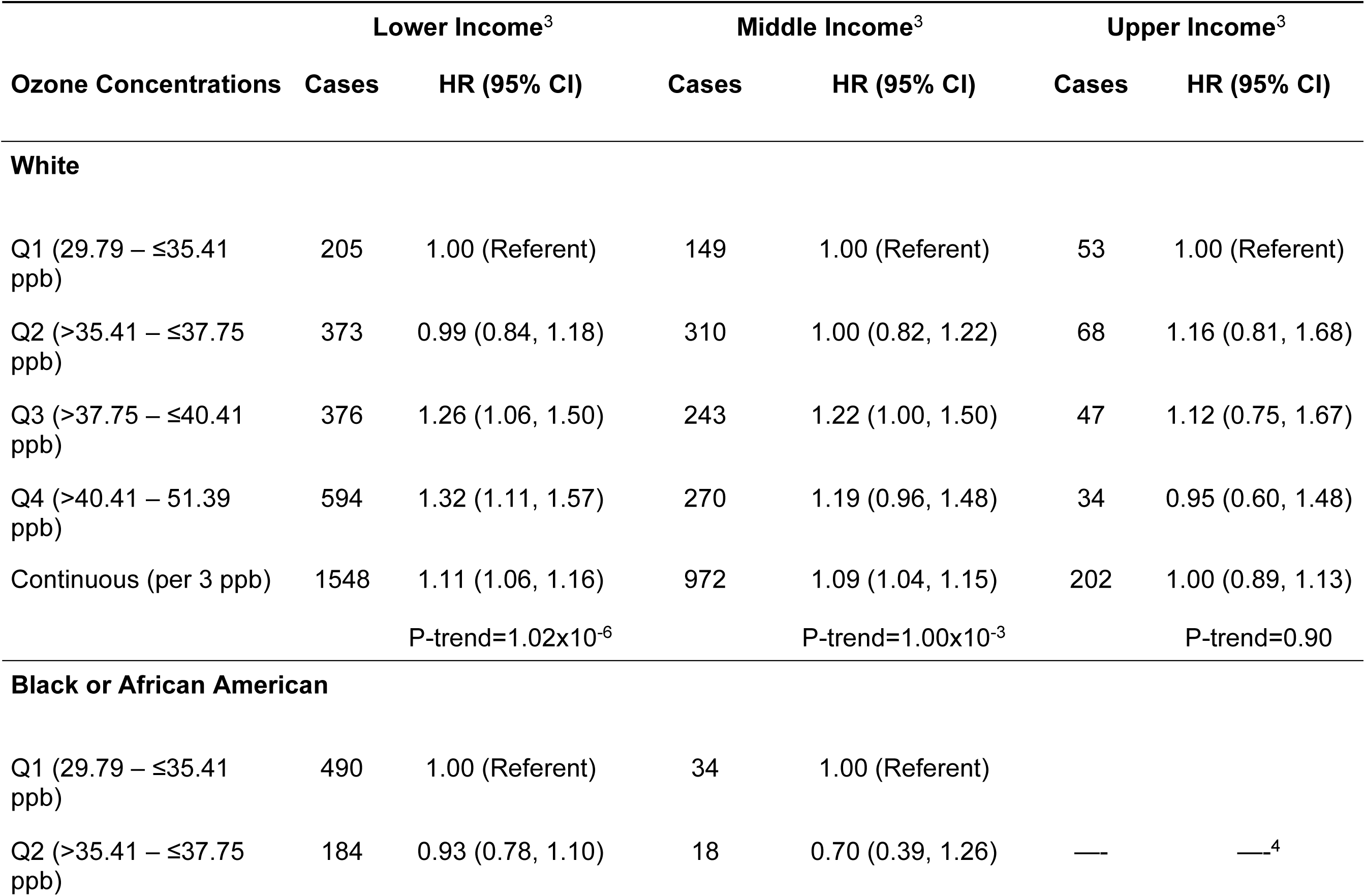
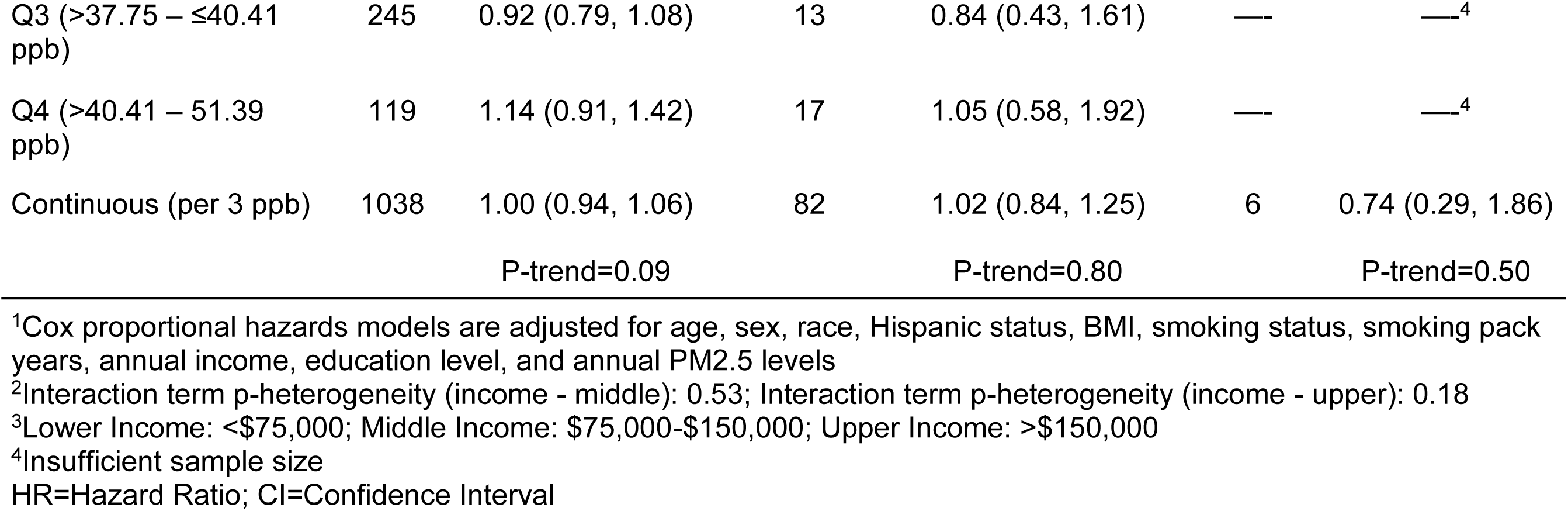
Associations between residential ground-level ozone concentrations and incident COPD risk by annual household income levels and race in the All of Us research program^1, 2^.

**Supplementary Table 2:** Associations between residential ground-level ozone concentrations and incident COPD risk by annual household income levels and sex in the All of Us research program^1, 2^.

| Lower Income<sup>3</sup> | Middle Income<sup>3</sup> | Upper Income<sup>3</sup> |
| --- | --- | --- |

| Ozone Concentrations | Cases | HR (95% CI) | Cases | HR (95% CI) | Cases | HR (95% CI) |
| --- | --- | --- | --- | --- | --- | --- |
| **Male** |  |  |  |  |  |  |
| Q1 (29.79 – ≤35.41 ppb) | 427 | 1.00 (Referent) | 102 | 1.00 (Referent) | 35 | 1.00 (Referent) |
| Q2 (>35.41 – ≤37.75 ppb) | 229 | 0.89 (0.75, 1.06) | 170 | 1.03 (0.80, 1.32) | 41 | 1.19 (0.75, 1.89) |
| Q3 (>37.75 – ≤40.41 ppb) | 286 | 1.04 (0.89, 1.21) | 151 | 1.36 (1.05, 1.75) | 30 | 1.15 (0.70, 1.89) |
| Q4 (>40.41 – 51.39 ppb) | 412 | 1.17 (1.00, 1.39) | 140 | 1.13 (0.87, 1.48) | 25 | 1.15 (0.68, 1.95) |
| Continuous (per 3 ppb) | 1354 | 1.06 (1.02, 1.11) | 563 | 1.07 (1.00, 1.14) | 131 | 1.03 (0.90, 1.19) |
|  |  | P-trend=0.01 |  | P-trend=0.06 |  | P-trend=0.60 |
| **Female** |  |  |  |  |  |  |
| Q1 (29.79 – ≤35.41 ppb) | 458 | 1.00 (Referent) | 104 | 1.00 (Referent) | 31 | 1.00 (Referent) |
| Q2 (>35.41 – ≤37.75 ppb) | 374 | 0.85 (0.74, 0.98) | 173 | 0.92 (0.71, 1.17) | 29 | 0.81 (0.49, 1.36) |
| Q3 (>37.75 – ≤40.41 ppb) | 385 | 0.98 (0.85, 1.13) | 128 | 1.06 (0.82, 1.38) | 22 | 0.83 (0.48, 1.43) |
| Q4 (>40.41 – 51.39 ppb) | 445 | 1.06 (0.90, 1.24) | 178 | 1.17 (0.90, 1.53) | 12 | 0.49 (0.24, 1.01) |
| Continuous (per 3 ppb) | 1662 | 1.03 (0.99, 1.07) | 583 | 1.10 (1.03, 1.18) | 94 | 0.88 (0.74, 1.06) |
|  |  | P-trend=0.20 |  | P-trend=0.01 |  | P-trend=0.20 |

<sup>1</sup>Cox proportional hazards models are adjusted for age, sex, race, Hispanic status, BMI, smoking status, smoking pack years, annual income, education level, and annual PM2.5 levels
<sup>2</sup>Interaction term p-heterogeneity (income - middle): 0.53; Interaction term p-heterogeneity (income - upper): 0.18
<sup>3</sup>Lower Income: <\$75,000; Middle Income: \$75,000-\$150,000; Upper Income: >\$150,000
HR=Hazard Ratio; CI=Confidence Interval

**Supplementary Table 3:** Associations between residential ground-level ozone concentrations and incident COPD risk accounting for temperature and nitrogen dioxide in the All of Us research program^1^.

| Ground-level ozone and Average Temperature Levels | Cases | HR (95%CI) |
| --- | --- | --- |
| Low Levels of ozone and Low Levels of Annual Average Temperature | 1327 | Reference Level |
| Low Levels of ozone and High Levels of Annual Average Temperature | 574 | 1.31 (1.18, 1.45) |
| High Levels of ozone and Low Levels of Annual Average Temperature | 374 | 1.20 (1.06, 1.35) |
| High Levels of ozone and High Levels of Annual Average Temperature | 1346 | 1.21 (1.11, 1.30) |

<sup>1</sup>Cox proportional hazards models are adjusted for age, sex, race, Hispanic status, BMI, smoking status, smoking pack years, annual income, education level, annual PM2.5 levels, and NO2 levels HR=Hazard Ratio; CI=Confidence Interval

**Supplementary Table 4:** Associations between residential ground-level ozone concentrations and incident COPD risk accounting for potential selection bias in the All of Us research program^1^.

| Ozone Concentrations | Cases | Overall (Unweighted) | State, age, race,<br>Hispanic status,<br>education, income | State, age, race, Hispanic<br>status, education, income,<br>marital status, military,<br>insurance, US born |
| --- | --- | --- | --- | --- |
|  |  | HR (95% CI) | HR (95% CI) | HR (95% CI) |
| Q1 (29.79 – ≤35.41 ppb) | 1551 | 1.00 (Referent) | 1.00 (Referent) | 1.00 (Referent) |
| Q2 (>35.41 – ≤37.75 ppb) | 1204 | 0.88 (0.81, 0.95) | 0.95 (0.84, 1.08) | 0.97 (0.85, 1.12) |
| Q3 (>37.75 – ≤40.41 ppb) | 1251 | 1.02 (0.95, 1.11) | 1.13 (1.00, 1.28) | 1.17 (1.02, 1.34) |
| Q4 (>40.41 – 51.39 ppb) | 1534 | 1.09 (1.01, 1.18) | 1.16 (1.03, 1.31) | 1.19 (1.03, 1.37) |
| Continuous (per 3 ppb) | 5540 | 1.10 (1.07, 1.13) | 1.10 (1.06, 1.15) | 1.13 (1.08, 1.18) |
|  |  | P-trend=6.59x10<sup>-13</sup> | P-trend=6.61x10<sup>-7</sup> | P-trend=1.00x10<sup>-7</sup> |

<sup>1</sup>Cox proportional hazards models are adjusted for age, sex, race, Hispanic status, BMI, smoking status, smoking pack years, annual income, education level, and annual PM2.5 levels
HR=Hazard Ratio; CI=Confidence Interval

